# HEALTH OUTCOMES IN COPD SMOKERS USING HEATED TOBACCO PRODUCTS: A 3-YEAR FOLLOW-UP

**DOI:** 10.1101/2020.12.10.20243857

**Authors:** R Polosa, JB Morjaria, U Prosperini, B Busà, A Pennisi, G Gussoni, S Rust, M Maglia, P Caponnetto

## Abstract

**Background:** Given that many patients with chronic obstructive pulmonary disease (COPD) smoke despite their symptoms, it is important to understand the long term health impact of cigarette substitution with heated tobacco products (HTPs). We monitored health parameters for 3-years in COPD patients who substantially attenuated or ceased cigarette consumption after switching to HTPs.

**Methods:** Changes in daily cigarette smoking, annualized disease exacerbations, lung function indices, patients reported outcomes (CAT scores) and 6-minute walk distance (6MWD) from baseline were measured in COPD patients using HTPs at 12, 24 and 36 months. These were compared to a group of age- and sex-matched COPD patients who continued smoking.

**Results:** Complete data sets were available for 38 patients (19 in each group). Subjects using HTPs had a substantial decrease in annualized COPD exacerbations within the group mean (±SD) from 2.1 (±0.9) at baseline to 1.4 (±0.8), 1.2 (±0.8) and 1.3 (±0.8) at 12-, 24- and 36-month follow-up (p<0.05 for all visits). In addition, substantial and clinically significant improvements in CAT scores and 6MWD were identified at all 3 time points in the HTP cohort. No significant changes were observed in COPD patients who continued smoking.

**Conclusions:** This study is the first to describe the long-term health effects of HTP use in COPD patients. Consistent improvements in respiratory symptoms, exercise tolerance, quality of life, and rate of disease exacerbations were observed in patients with COPD who abstained from smoking or substantially reduced their cigarette consumption by switching to HTP use.

## INTRODUCTION

Chronic obstructive pulmonary disease (COPD) is an umbrella term used to describe distinct obstructive lung conditions including chronic bronchitis and emphysema (1,2). With over 3 million deaths, COPD is the world’s fourth leading cause of death at present (3). Individuals experience the ill effects of this disease for years, and die prematurely from it or its cardio-respiratory complications (4,5).

Globally, the COPD burden is expected to increase in the coming decades due to continued exposure to COPD risk factors and/or population ageing (6).

It is well known that cigarette smoking is a significant risk factor for COPD (7-9) and abstaining from smoking is the only evidence

based intervention that improves the prognosis for COPD (10,11)

Although stopping smoking should be a priority for any smokers with COPD, most of them are unable to experience high success rates during their quit attempts (12,13). Licensed quitting therapies (i.e. nicotine replacement therapy (NRT), bupropion, and varenicline) have only limited success in terms of a sustained cessation long-term in patients with COPD who smoke (14).

These patients struggle to completely stop nicotine use, and may require prolonged treatment and/or sustained nicotine use to achieve longstanding abstinence from smoking.

An alternative pragmatic approach for patients with COPD who are having difficulty at stopping smoking is that of substituting conventional cigarettes with combustion-free nicotine delivery alternatives to achieve significant health gains (15,16).

Although not risk free, emerging clinical evidence suggests that, for example, e-cigarettes use in patients with COPD can help patients with COPD abstain from long-term smoking with clinically relevant health gains (17,18).

More recently, another class of combustion-free products has been introduced for cigarette substitution, which is also gaining popularity and acceptance by consumers worldwide (19-21). Heated tobacco products, (HTP, also known as heat-not-burn), consist of a holder that electronically transfers controlled heat at temperatures which are below 350°C (instead of burning tobacco) to tobacco sticks, plugs, or capsules that generate a nicotine-containing aerosols.

Since these aerosols are produced at much lower temperatures (compared to combustion, which generally starts at temperature above 400°C), they contain less harmful and potentially harmful chemicals than tobacco smoke, and the overall level of chemical exposure has been shown to be significantly lower in exclusive HTP users compared to smokers (22,23). Most popular HTPs are commercially known as “IQOS” by Philip Morris International, “glo” by British American Tobacco, and “Ploom TECH” by Japan Tobacco International. In July 2020, the U.S. FDA authorized the marketing of Philip Morris’s “IQOS Tobacco Heating System” as modified risk tobacco products (MRTPs), the first tobacco product to be legally commercialized with the claim of reducing consumers’ exposure to harmful chemicals when completely switching away from conventional cigarettes (24).

Given that HTPs have been shown to generate substantially less toxic and potentially harmful chemicals than conventional cigarettes, it is hypothesized that when substituting combustible tobacco cigarettes for HTPs, clinically important health improvements can be observed.

Knowledge of the long-term health impacts of HTP use in this patient population is needed to counsel COPD patients who are using or intend to use HTPs.

This awareness is very limited and we are unaware of any published research that has reported the long-term health consequences of the use of HTPs in COPD. This study is the first to assess objective and subjective health parameters in a cohort of COPD patients who have been daily users of HTPs. Findings were assessed over an observation period of 3-years and compared to those obtained from an age-, sex-matched cohort of smokers with COPD.

## METHODS

### Patient population

We conducted a review of medical records of COPD patients regularly attending outpatient clinics in four Italian hospitals. The diagnosis of COPD was made in accordance with the criteria set out by the Global Initiative for Chronic Obstructive Lung Disease (GOLD) (25).

Patients with COPD who reported using HTPs on at least 2 consecutive outpatient visits (no less th an 12 months apart) were deemed eligible and included in the study (HTP users cohort). Datasets from age- and sex-matched COPD patients who regularly smoked cigarettes (and were not using HTPs or e-cigarettes) over the same observation period and attending the same clinics were included as a reference group (cigarette smokers cohort). Approval for the study was obtained by the coordinating center’s ethics review board and each patient provided written informed consent.

### Study design and Assessments

Study design and evaluations were previously described (26,27). For the cohort of HTP users, the baseline visit was considered a clinical visit prior to the first of the two consecutive follow-up visits, when HTPs were not yet used by COPD patients and they were smoking cigarettes. HTP devices have been noted. Prospective data from COPD HTP users and COPD cigarette smokers were collected at 12±1.5 (Follow-up visit : F/up1)) 24 ±1.5 (F/up2) and 36 ±2.5 (F/up3) months from October 2017 to September 2020 during their annual follow-up visits.

Clinical, behavioural, and functional parameters were compared between study cohorts and included: (i) respiratory symptoms, (ii) smoking status (biochemically confirmed by exhaled breath carbon monoxide - eCO), cigarette consumption per day (cig/day), as well as HTP use, (iii) the annual number of severe COPD exacerbations, (iv) post-bronchodilator lung function parameters (forced expiratory flow in 1 second - FEV1; forced vital capacity - FVC; expiratory ratio - FEV1/FVC; (v) patients’ reported outcomes (by COPD Assessment Test – CAT scores) and (vi) level of exercise tolerance (by 6 Minute Walking Distance - 6MWD test). Additionally, we also assessed variations in the relative proportion of COPD GOLD stages over the 3-year study period. The study objectives were not known to the hospital staff who carried out data extraction from patients’ medical records. Anonymized datasets were analyzed by a statistician who was blinded to the study cohorts.

Severe exacerbations were defined as changes in the patients’ respiratory symptoms requiring a course of antibiotics and/or systemic corticosteroids in addition to nebulized bronchodilators via the primary care physician, emergency department attendance, and/or hospital admission.

The CAT is a health status validated questionnaire used in COPD patients. A 2 unit difference is considered a minimal clinically important difference in patients’ symptomatology (28,29). The 6MWD is an assessment of the general ability of patients to perform daily tasks. Only patients who were physically capable of carrying out the test underwent 6MWD (30).

### Smoking and HTP status

Smoking consumption was recorded at each study visit as the number of cigarettes smoked on the day before the visit. Smoking abstinence was described as a complete self-reported cessation of conventional smoking (not even a puff) since the previous study visit and biochemically verified by an eCO level of ⍰7 ppm. COPD HTP users who completely stopped cigarette smoking were described as quitters (single users), whereas patients who reported using HTP in conjunction with conventional cigarette smoking were defined as dual users.

### Data management and statistical analyses

Patients’ demographic and clinical data were noted at each outpatient visit. Patient data was collected and imputed into an electronic spreadsheet before statistical analyses. Of note, the investigators involved in the analyses of the data did not participate in the medical supervision of the participants in the study or in the extraction of data from the medical records.

In the current analyses, patient parameters are presented as means [± standard deviation (SD)] and medians [interquartile range (IQR)] for parametric and non-parametric data, respectively. Differences in baseline data between the two groups were conducted using student’s **t**-test and Mann Whitney U Test for parametric and non-parametric data. Data from single and dual users were also extracted for secondary analyses. Statistical within group analyses using the student’s t-test and Wilcoxon-signed rank test were conducted for parametric or non-parametric data at the 3 time points compared to baseline, respectively. Analysis of repeated measures with Bonferroni correction between the two study groups was conducted for repeated parameter measurements over the 3-years. A two-tailed p-value of less than 0.05 was considered of statistical significance. The Statistical Package for Social Science (SPSS for Windows, version 20.0, Chicago, IL, USA) was used to conduct all the statistical assessments.

## RESULTS

### Patient characteristics

A total of 44 COPD patients were considered for the study at baseline, but complete datasets were available for analyses from 38 COPD patients (31 male, 7 female) by the end of the study. Three COPD patients in the HTP user group discontinued the use of their device during follow-up (two resulting in relapse to cigarette smoking, one stopping using tobacco products completely) and were excluded. Datasets from three COPD patients of the control group were not available for analysis (one moved to a different city area, and two transferred to stroke clinics). The baseline demographics, parameters assessed and COPD GOLD staging are outlined on **Table 1**. There were no statistical differences between the 2 study groups for any of the parameters. Most patients had mild-to-severe airflow limitation as per the COPD GOLD guidelines, and were managed accordingly with various combinations of inhaled therapies (corticosteroids, β2 agonists and long-acting anti-cholinergics, individually or in combination).

**Table 1.**
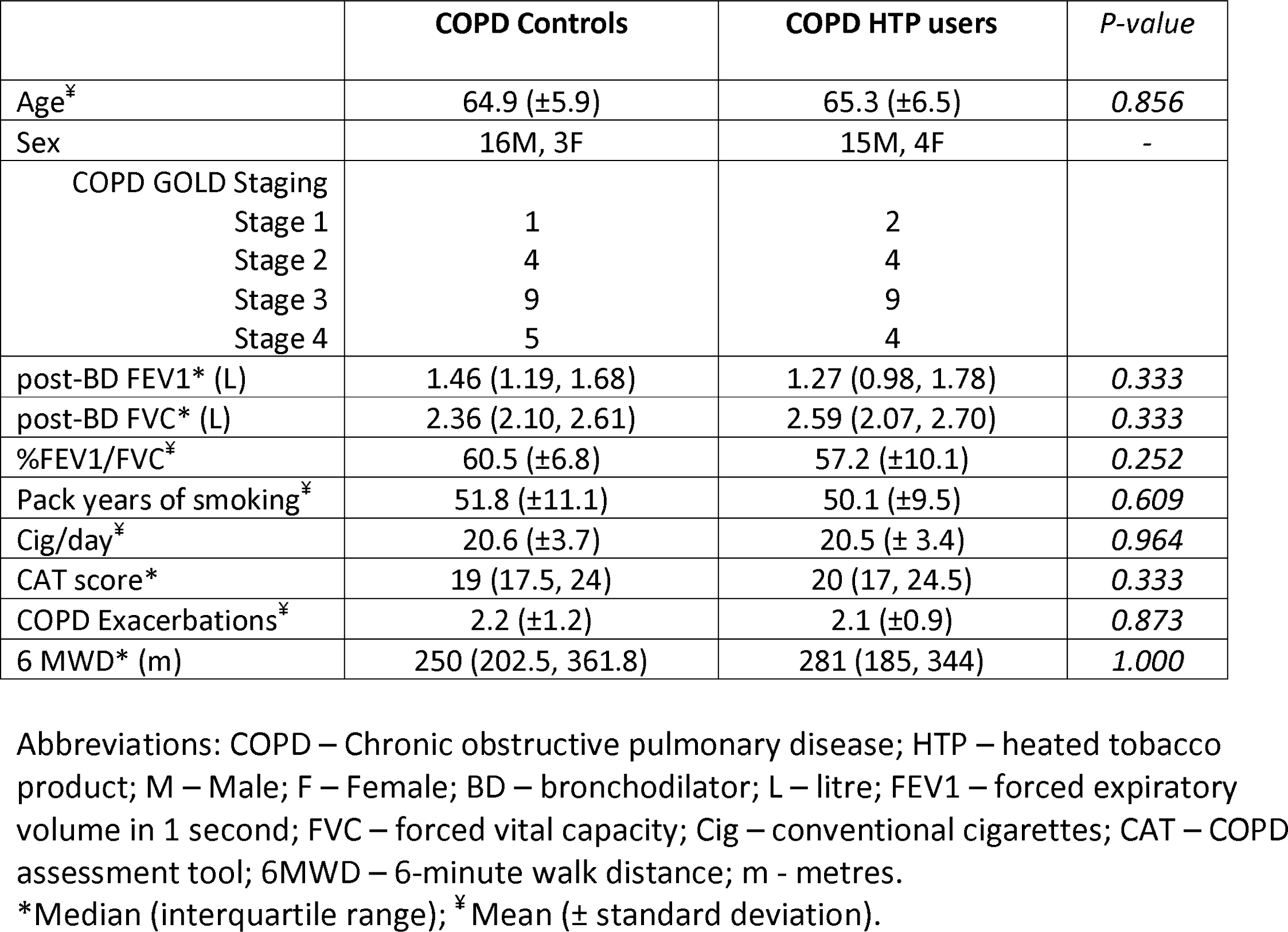
Baseline demographics of the subjects on the study.

|  | **COPD Controls** | **COPD HTP users** | *P-value* |
| --- | --- | --- | --- |
| Age<sup>‡</sup> | 64.9 (±5.9) | 65.3 (±6.5) | 0.856 |
| Sex | 16M, 3F | 15M, 4F | - |
| COPD GOLD Staging |  |  |  |
| Stage 1 | 1 | 2 |  |
| Stage 2 | 4 | 4 |  |
| Stage 3 | 9 | 9 |  |
| Stage 4 | 5 | 4 |  |
| post-BD FEV1* (L) | 1.46 (1.19, 1.68) | 1.27 (0.98, 1.78) | 0.333 |
| post-BD FVC* (L) | 2.36 (2.10, 2.61) | 2.59 (2.07, 2.70) | 0.333 |
| %FEV1/FVC<sup>‡</sup> | 60.5 (±6.8) | 57.2 (±10.1) | 0.252 |
| Pack years of smoking<sup>‡</sup> | 51.8 (±11.1) | 50.1 (±9.5) | 0.609 |
| Cig/day<sup>‡</sup> | 20.6 (±3.7) | 20.5 (± 3.4) | 0.964 |
| CAT score* | 19 (17.5, 24) | 20 (17, 24.5) | 0.333 |
| COPD Exacerbations<sup>‡</sup> | 2.2 (±1.2) | 2.1 (±0.9) | 0.873 |
| 6 MWD* (m) | 250 (202.5, 361.8) | 281 (185, 344) | 1.000 |

Abbreviations: COPD – Chronic obstructive pulmonary disease; HTP – heated tobacco product; M – Male; F – Female; BD – bronchodilator; L – litre; FEV1 – forced expiratory volume in 1 second; FVC – forced vital capacity; Cig – conventional cigarettes; CAT – COPD assessment tool; 6MWD – 6-minute walk distance; m - metres.
\*Median (interquartile range); <sup>‡</sup>Mean (± standard deviation).

### Cigarette Consumption and HTP use

In the COPD HTP users, a significant reduction in conventional cigarette use was noted with a mean (±SD) cigarettes/day of 20.5 (±3.4) at baseline falling to 1.5 (±2), 1.2 (±1.8) and 1.2 (±1.8) at F/up1, F/up 2, and F/up 3, respectively (p<0.001 for all 3 visits) (**Table 2; Figure 1**). There were no remarkable differences over the course of the study in the number of cigarettes smoked per day in the COPD controls. Among COPD HTP users, 11/19 (57.9%) completely abstained from smoking conventional cigarettes (*exclusive HTP users/single users*) at F/up 3 (**Table 3**). In those continuing to smoke (*dual users*) there was a considerable decline in daily cigarette consumption, the mean (±SD) cigarettes/day at baseline decreasing from 21 (±4.6) to 3.6 (±1.1), 3.3 (±1.3) and to 3.7 (±1) at F/up1, F/up2 and F/up3, respectively (p<0.001 for all three visits) (**Table 3**). Of note, all dual users consistently reduced their daily smoking by at least 70% of their baseline consumption throughout the whole duration of the study. Overall, there was a marked reduction in daily cigarettes smoked between the two study groups over the 36-month observation period (p<0.001).

**Table 2.** Comparison of controls and HTP users at baseline, 12-, 24- and 36-month follow-up visits.

|  | Baseline | 12-Month<br>Follow-up | P value<sup>Φ</sup> | 24-Month<br>Follow-up | P value<sup>Φ</sup> | 36-Month<br>Follow-up | P value<sup>Φ</sup> | *Overall*<br>*between*<br>*group p*<br>*value from*<br>*Baseline* |
| --- | --- | --- | --- | --- | --- | --- | --- | --- |
| **COPD Controls**<br>**(n=19)** |  |  |  |  |  |  |  |  |
| post-BD FEV1* (L) | 1.46 (1.19, 1.68) | 1.41 (1.17, 1.68) | 0.614 | 1.36 (1.18, 1.67) | 0.872 | 1.47 (1.18, 1.62) | 0.643 | *0.469* |
| post-BD FVC* (L) | 2.36 (2.1, 2.61) | 2.27 (2.2, 2.71) | 0.135 | 2.31 (2.16, 2.81) | 0.251 | 2.3 (2.08, 2.57) | 0.149 | *0.385* |
| %FEV1/FVC<sup>‡</sup> | 60.5 (±6.8) | 59.4 (±6.7) | 0.631 | 59.2 (±6.8) | 0.575 | 60.8 (±8.7) | 0.594 | *0.239* |
| Cig/day<sup>‡</sup> | 20.6 (±3.7) | 20.2 (±3.8) | 0.764 | 19.8 (±5.0) | 0.657 | 19.6 (±4) | 0.650 | **<0.001** |
| CAT score* | 19 (17.5, 24) | 20 (18, 22) | 0.284 | 20 (15.5, 24.5) | 0.520 | 20 (18, 23) | 0.709 | **0.008** |
| COPD<br>Exacerbations<sup>‡</sup> | 2.2 (±1.1) | 2.2 (±1) | 0.878 | 2.1 (±1.1) | 0.833 | 2.1 (±0.9) | 0.728 | **0.024** |
| 6MWD*<sup>Σ</sup> | 250 (202.5, 361.8) | 270 (210.5, 368.8) | 0.872 | 263.5 (225.5, 374.8) | 0.182 | 270 (216, 362) | 0.155 | **0.001** |
| **COPD HTP users**<br>**(n=19)** |  |  |  |  |  |  |  |  |
| post-BD FEV1* (L) | 1.27 (0.98, 1.78) | 1.33 (0.99, 1.77) | 0.120 | 1.39 (1.05, 1.72) | 0.064 | 1.30 (1.02, 1.79) | 0.257 |  |
| post-BD FVC* (L) | 2.59 (2.07, 2.7) | 2.6 (2.07, 2.76) | 0.277 | 2.57 (2.02, 2.91) | 0.296 | 2.55 (2.03, 2.93) | 0.376 |  |
| %FEV1/FVC<sup>‡</sup> | 57.2 (±10.1) | 57.1 (±10.5) | 0.980 | 58.1 (±10.2) | 0.798 | 57.3 (±10.3) | 0.984 |  |
| Cig/day<sup>‡</sup> | 20.5 (± 3.4) | 1.5 (±2) | **<0.001** | 1.2 (±1.8) | **<0.001** | 1.2 (±1.8) | **<0.001** |  |
| CAT score* | 20 (17, 24.5) | 16 (14.5, 20) | **<0.001** | 17 (14.5, 19) | **0.001** | 15 (13, 21) | **0.006** |  |
| COPD<br>Exacerbations<sup>‡</sup> | 2.1 (±0.9) | 1.4 (±0.8) | **0.012** | 1.2 (±0.8) | **0.002** | 1.3 (±0.8) | **0.004** |  |
| 6MWD*<sup>Σ</sup> (m) | 281 (185, 344) | 310 (219.5, 370) | **0.004** | 333 (224, 370) | **0.003** | 350 (249, 396) | **0.005** |  |

Abbreviations: COPD – Chronic obstructive pulmonary disease; HTP – heated tobacco product; n – number; BD – bronchodilator; L – litre; FEV1 – forced expiratory volume in 1 second; FVC – forced vital capacity; Cig – conventional cigarettes; CAT – COPD assessment tool; 6MWD – 6 minute walk distance; m – metres.
\*Median (interquartile range); ‡ Mean (± standard deviation).
Φ within group P value vs Baseline
Σ 11 subjects in the COPD HTP user group and 11 in the COPD control group at all 3 follow-up visits.
Significant differences are noted in bold.

**Table 3.** Comparison of HTP and conventional cigarette users (dual users) vs HTP exclusive users (single users) at 12-, 24- and 36-month.

| Parameter | Baseline | 12-Month Follow-up | *p value*<sup>Φ</sup> | 24-Month Follow-up | *p value*<sup>Φ</sup> | 36-Month Follow-up | *p value*<sup>Φ</sup> |
| --- | --- | --- | --- | --- | --- | --- | --- |
| **COPD HTP users reducing cig use (dual users)** | **(n=8)** | **(n=8)** |  | **(n=7)** |  | **(n=6)** |  |
| Sex | 7M,1F | 7M,1F |  | 7M |  | 6M |  |
| % Smoking reduction from baseline | - | 82 (±5.7)% | - | 83.9 (±6.2)% | - | 81.8 (±5.9)% | - |
| post-BD FEV1* (L) | 1.27 (0.97, 1.88) | 1.37 (0.99, 2.01) | **0.024** | 1.85 (0.99, 2.13) | 0.063 | 1.53 (0.99, 1.97) | 0.686 |
| post-BD FVC* (L) | 2.47 (2.03, 3.05) | 2.58 (2.13, 3.1) | **0.050** | 2.91 (2.37, 3.12) | 0.176 | 2.84 (2.35, 3.12) | 1.000 |
| %FEV/FVC<sup>‡</sup> | 56.1 (±9.1) | 55.9 (±10.3) | 0.980 | 57 (±12.1) | 0.863 | 53.7 (±10.8) | 0.873 |
| Cig/day<sup>‡</sup> | 21 (±4.6) | 3.6 (±1.1) | **<0.001** | 3.3 (±1.3) | **<0.001** | 3.7 (±1) | **<0.001** |
| CAT score* | 23 (18.3, 26) | 20 (16, 21) | **0.018** | 18 (13.5, 21) | **0.027** | 19 (13.5, 23.8) | **0.027** |
| COPD Exacerbations<sup>‡</sup> | 2.1 (±0.6) | 1.6 (±0.5) | **0.108** | 1.1 (±0.7) | 0.077 | 1.2 (±0.4) | **0.021** |
| **COPD HTP users ceasing cig use (single users)** | **(n=11)** | **(n=11)** |  | **(n=12)** |  | **(n=13)** |  |
| Sex | 8M, 3F | 8M, 3F |  | 8M, 4F |  | 9M, 4F |  |
| % Smoking reduction from baseline | - | 100% | - | 100% | - | 100% | - |
| post-BD FEV1* (L) | 1.38 (1.03, 1.71) | 1.26 (1.02, 1.75) | 0.722 | 1.35 (1.17, 1.64) | 0.504 | 1.3 (1.1, 1.65) | 0.401 |
| post-BD FVC* (L) | 2.6 (2.19, 2.67) | 2.6 (2.7, 2.73) | 0.689 | 2.44 (1.83, 2.83) | 0.814 | 2.5 (1.8, 2.78) | 0.727 |
| %FEV/FVC<sup>‡</sup> | 58.1 (±11) | 58 (±11) | 0.991 | 58.7 (±9.2) | 0.675 | 59 (±10) | 0.881 |
| Cig/day<sup>‡</sup> | 20.2 (±2.3) | - |  | - |  | - |  |
| CAT score* | 18 (17, 23) | 15 (14, 17) | **0.004** | 16 (14.8, 18) | **0.003** | 15 (14, 20) | **0.036** |
| COPD Exacerbations<sup>‡</sup> | 2.1 (±1) | 1.2 (±1) | **0.048** | 1.3 (±0.9) | **0.021** | 1.3 (±1) | **0.035** |

Abbreviations: n – number; COPD – Chronic obstructive pulmonary disease; HTP- heated tobacco product; M – male; F – female; BD – bronchodilator; L – litre; FEV1 – forced expiratory volume in 1 second; FVC – forced vital capacity; cig – conventional cigarettes; CAT – COPD assessment tool.
\*Median (interquartile range); <sup>‡</sup> Mean (± standard deviation).
$\Phi$ within group P value vs Baseline
Significant differences are noted in bold.

**Figure 1.**
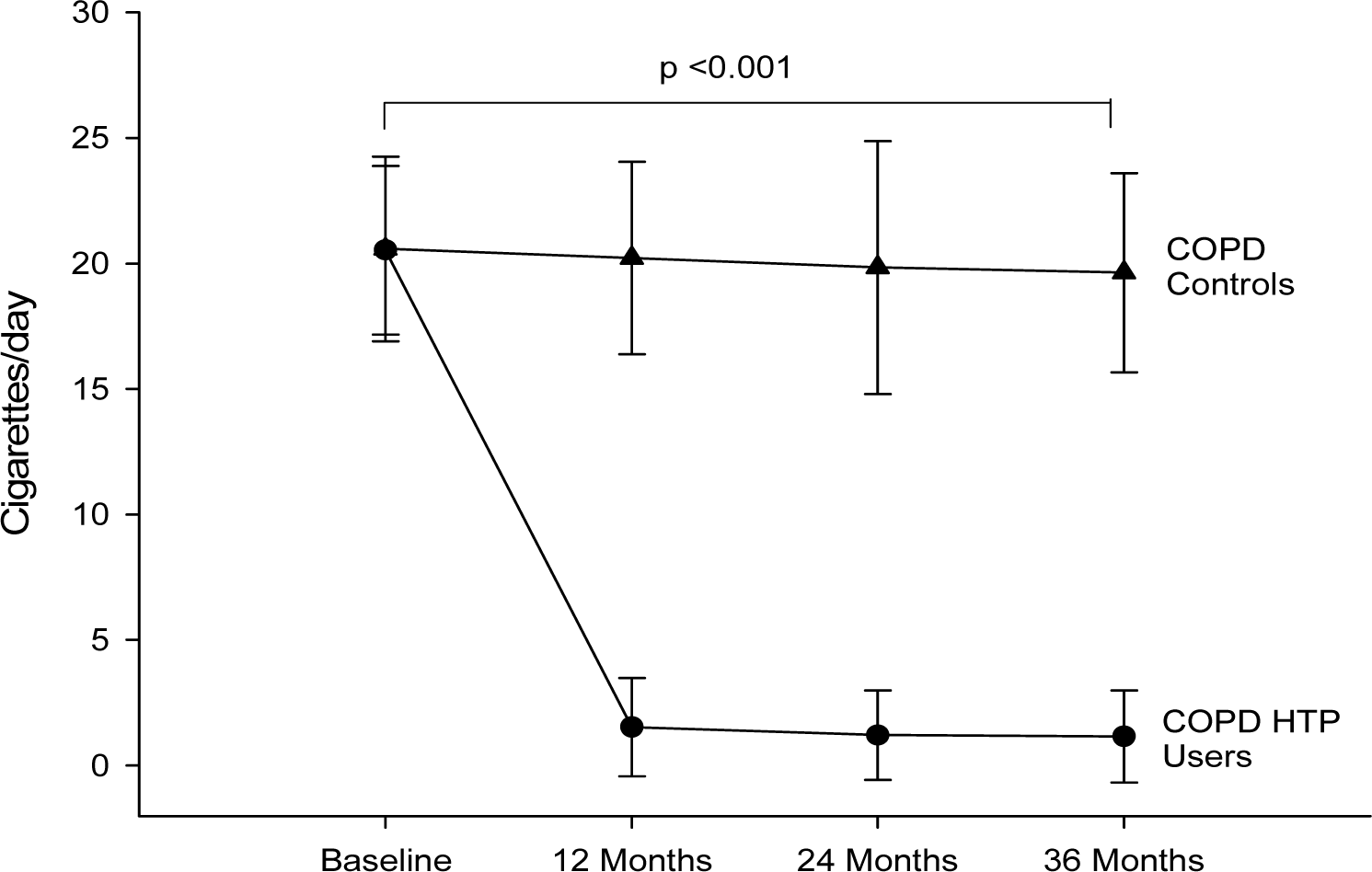
Number of cigarettes smoked per day at baseline, follow-up visit 1 (12 ±1.5 months), visit 2 (24 ±2.5 months) and visit 3 (36 ±3 months) in COPD heated tobacco product users (closed circles) and COPD controls (closed triangles). All data expressed as mean and error bars are standard deviation of the mean. *Abbreviations:* COPD: chronic obstructive pulmonary disease; HTP: heated tobacco products.

At F/up1, all HTP users were on IQOS. Three IQOS users were found to have switched to glo at F/up2. At F/up3, 17 HTP users were on IQOS and two on glo. No detail on tobacco sticks consumption was recorded.

### COPD Exacerbations

COPD HTP users had a significant decline in COPD exacerbations; with the mean (±SD) annual exacerbation rate decreasing from 2.2 (±1.1) at baseline to 1.4 (±0.8)(p=0.012), 1.2 (±0.8)(p=0.002) and 1.3 (±0.8) (p=0.004) at F/up1, F/up2 and F/up3, respectively (**Table 2**). No significant changes in the annual COPD exacerbation rates were observed in the control group. There was an overall significant (p=0.024) between group decrease in annual COPD exacerbations over observation period (**Table 2**; **Figure 2**). In the exclusive (single) HTP users significant reductions in annual COPD exacerbations from baseline were noted at all 3 F/up visits (**Table 3**). Of note, a steady decline in annual COPD exacerbations was also observed in dual users, the mean (±SD) yearly exacerbation rate of 2.1 (±0.6) at baseline declining to 1.6 (±0.5)(p=0.108) at F/up1, 1.1 (±0.7)(p=0.077) at F/up 2 and 1.2 (±0.4) at F/up3 (p=0.021) (**Table 3**).

**Figure 2.**
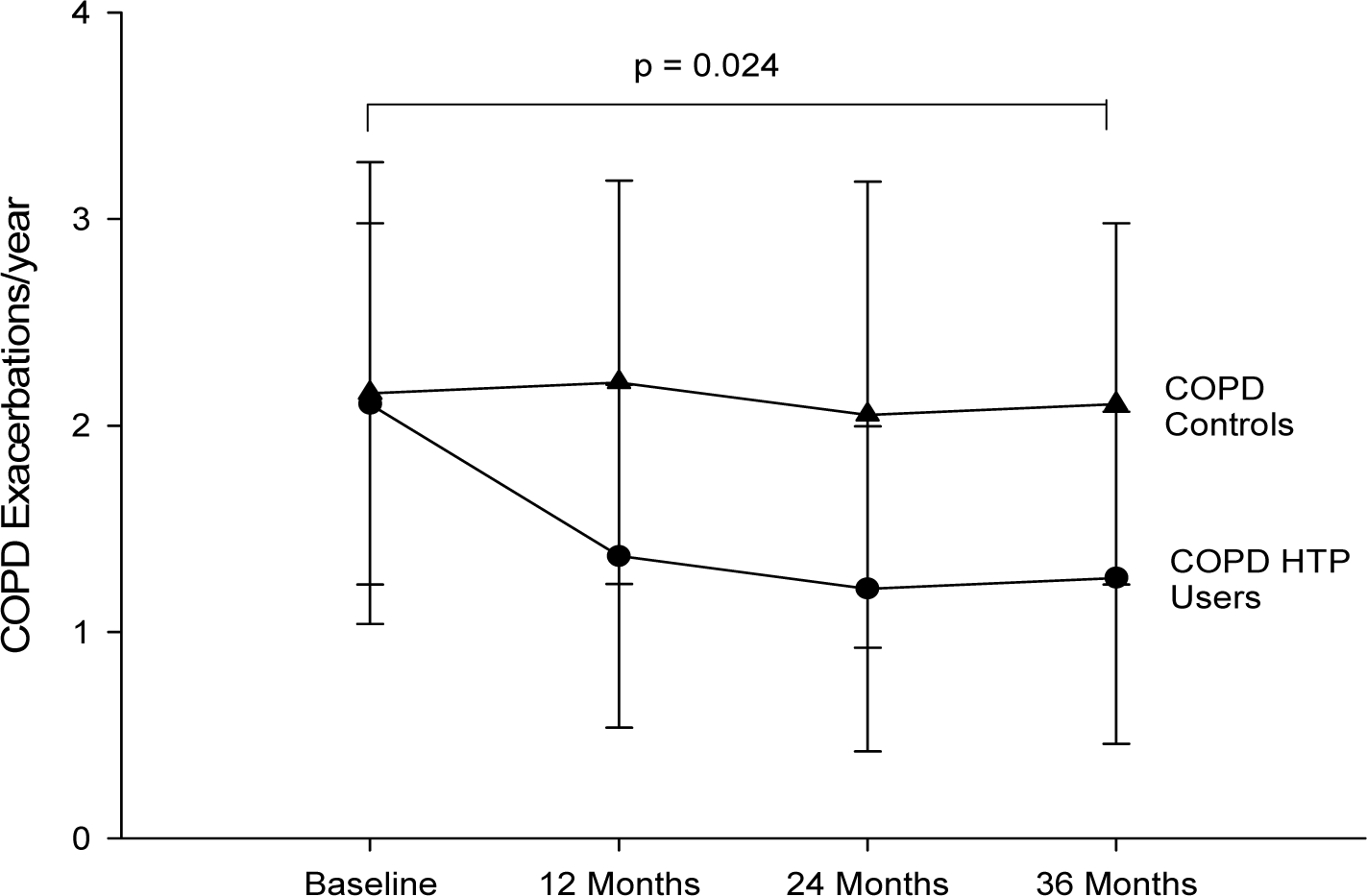
Number of COPD exacerbations per year at baseline, follow-up visit 1 (12 ±1.5 months), visit 2 (24 ±2.5 months) and visit 3 (36 ±3 months) in COPD heated tobacco product users (closed circles) and COPD controls (closed triangles). Data expressed as mean and error bars are standard deviation of the mean. *Abbreviations:* COPD: chronic obstructive pulmonary disease; HTP: heated tobacco products.

### Lung Function assessments and COPD staging

There was no significant post-baseline improvement in post bronchodilator FEV1 and FVC at any of the follow-up visits in the COPD HTP users (**Table 2**). Likewise, no significant change in spirometric indices were observed in the control group throughout the study (**Table 2**). Overall there were no significant differences between the two groups in the spirometric assessments (**Table 2**).

GOLD COPD staging changes throughout the study are illustrated in (**Table 4; Figure 3)**. By the end of the study, three COPD patients in the HTP user group down-staged (i.e. improved) from GOLD COPD Stages 4 and 3 to Stages 2 and 1; in contrast, COPD GOLD stage classification was relatively stable in patients belonging to the control group throughout the whole duration of the study.

**Table 4.**
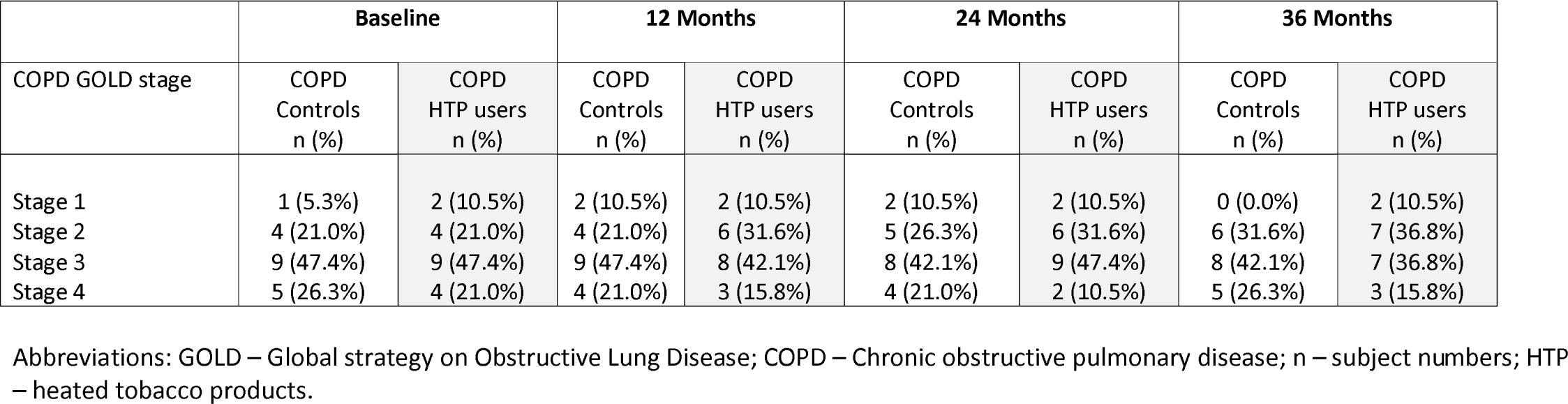
COPD GOLD stage changes over the study period.

**Figure 3.**
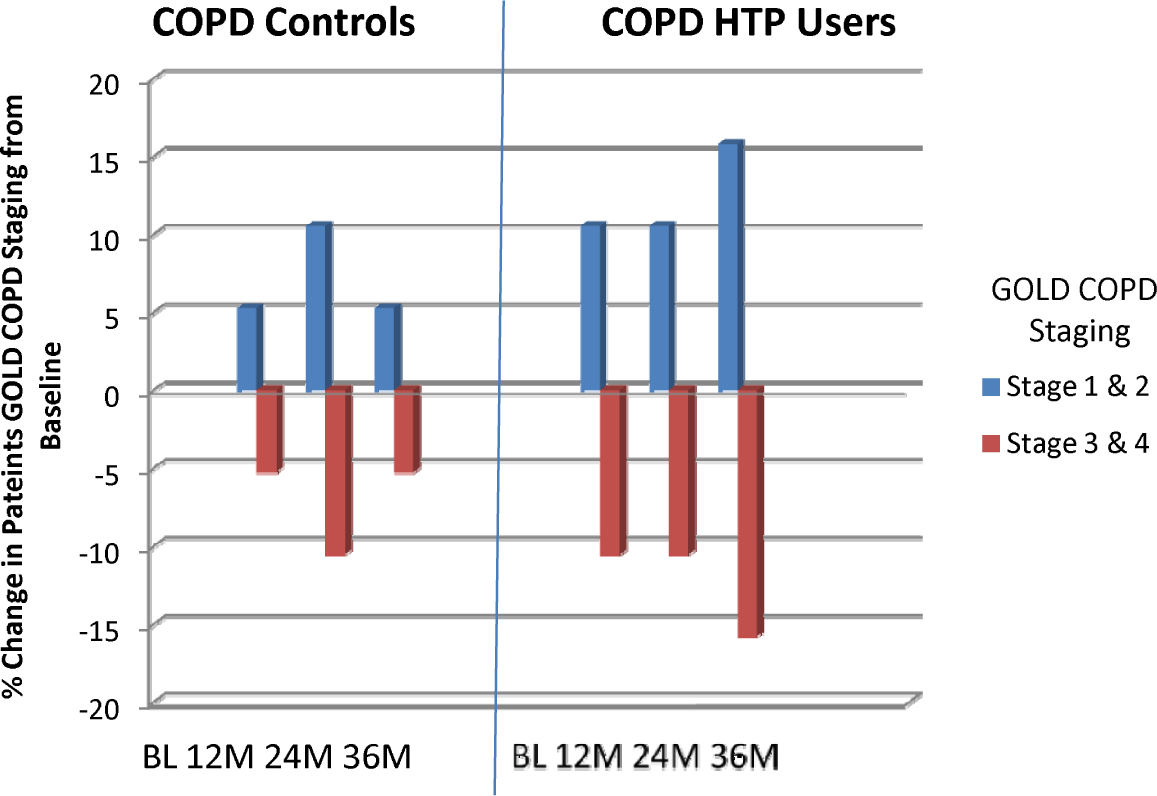
Percentage change in patients COPD GOLD stage from baseline in COPD heated tobacco product users and COPD controls. *Abbreviations:* COPD: chronic obstructive pulmonary disease; HTP: heated tobacco products; GOLD: Global initiative in Obstructive Lung Disease; BL: baseline; M: months.

### CAT scores and 6MWD test

Subjective COPD assessment using CAT scores significantly improved in the COPD HTP user group at all three follow-up visits compared to baseline (p<0.01 at all follow-up visits) (**Table 2**). Both dual and single users experienced significant reductions (improvements) in CAT scores from baseline (**Table 3**). In contrast there were no significant or clinically relevant improvements noted at any of the follow-up visits from baseline in the control group. Over the 3-year study period we observed an overall significant improvement in CAT scores between COPD HTP users and COPD smokers (p=0.008) (**Table 2**; **Figure 4**).

**Figure 4.**
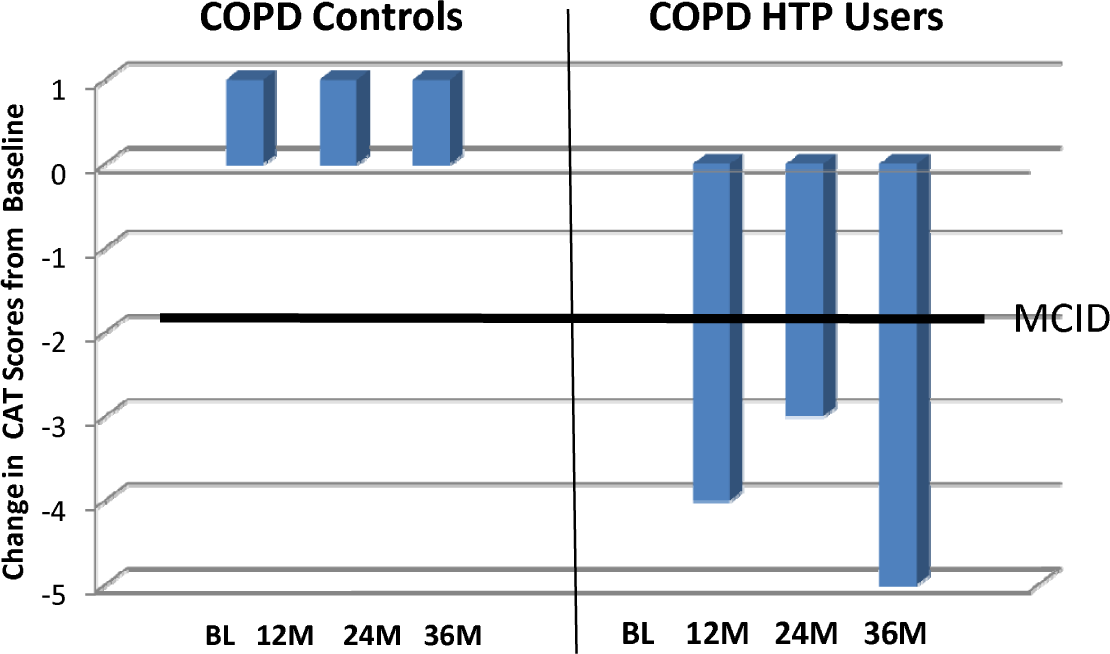
Change in the median COPD assessment tool (CAT) scores from baseline in COPD heated tobacco product users and COPD controls. The bold solid line on the bar chart represents the minimal clinically important difference (MCID) for CAT score. A decrease of at least 2 units from baseline is considered to be of clinical importance. *Abbreviations:* COPD: chronic obstructive pulmonary disease; HTP: heated tobacco products; CAT: COPD Assessment Tool; BL: baseline; M: Months; MCID: minimal clinically important difference.

Results of 6MWD at all four follow-up visits were available only for 22 patients (11 from each study group). In the HTP user group, the 6MWD significantly improved from baseline at all follow-up visits (p<0.01) whereas no remarkable improvements in 6MWD was observed in the control group (**Table 2**); at F/up3, we observed a median increment in 6MWD of 69m (p=0.005) in the COPD HTP user group whereas there was a small but not significant increase in median 6MWD of 20m (p=0.155) in the COPD control group (**Table 2**). Overall there was a significant improvement (p=0.001) in 6MWD noted between the two study groups over the entire follow-up period (**Table 2**).

## DISCUSSION

This study is the first to describe the long-term health effects of daily HTP use in COPD patients. Patients with COPD who abstained from smoking or substantially reduced their cigarette consumption by switching to HTP use experienced improvements in several objective and subjective health parameters, which persisted for up to 3 years. These findings with HTPs were not unexpected because avoiding exposure to chemicals generated from the combustion of tobacco cigarettes is known to slow COPD progression and to improve patients’ health (10, 31-33). Moreover, similar findings have been also reported in COPD patients using another class of combustion-free nicotine delivery products (i.e. e-cigarettes) (18, 26, 27).

Nearly 60% of COPD patients using HTPs abstained completely from cigarette smoking throughout the duration of the study, whereas those continuing to smoke (*dual users*) showed a consistent decline in their daily cigarette consumption from baseline of at least 70% at all study visits. This remarkable reduction in overall cigarette consumption may be explained by effective substitution of conventional cigarettes with HTPs in the COPD patients under investigation. By mimicking the experience of tobacco smoking and its related rituals, HTP use may provide adequate compensatory physical and behavioural effects (34,35), possibly serving as an effective relapse prevention method and therefore contributing to the low relapse rates observed in the HTP users of this study.

Holding off from chronic exposure to combustion chemicals is expected to improve several health outcomes in COPD patients, but long-term health consequences of regular HTP use have not been explored. An important finding is that COPD exacerbations were consistently reduced throughout the whole duration of the study by approximately 40% in patients who stopped or considerably reduced their smoking consumption after switching to HTPs. The proportion of COPD exacerbations prevented in these patients is of clinical significance and similar to that observed with standard medications (36). Chronic exposure to cigarette smoke enhance susceptibility to infections of the respiratory tract (37) and it has been shown to be a risk factor for bacterial and viral infections (38,39) as well as for respiratory exacerbations in COPD patients (40). Given that stopping smoking is associated with a lower risk, switching to HTP use would be expected to result in a marked attenuation of respiratory infections and COPD exacerbations, as shown in this study.

Consistent improvements were also observed in overall health status and physical activity in COPD patients who quit or reduced substantially their cigarette consumption by substitution with HTPs. The improvements in patients’ reported outcomes (by CAT scores) and level of exercise tolerance (by 6MWD test) in the COPD HTP user group were consistent throughout the whole duration of the study. Of note, significant improvements in CAT scores were observed for both dual users and exclusive HTP users. The observed CAT and 6MWD changes in this study are similar to those reported in COPD patients undergoing intensive rehabilitation programs (29,41). These improved health outcomes may be explained by the marked decline in carbon monoxide exposure and in carboxyhaemoglobin levels following cigarettes substitution with combustion-free HTPs (42) and the associated time-dependent improvement in exercise tolerance that occurs after quitting smoking (43). The same explanation may also hold up for dual users given that they greatly reduced their daily smoking consumption by more than 70%.

Given the improvements in exacerbation rates, respiratory symptoms, and overall health status, it was not surprising to note for COPD patients in the HTP user group, consistent downstaging (i.e. improvement) of their GOLD COPD stages throughout the whole duration of the study.

No significant post-baseline improvement in spirometric indices was observed in COPD HTP users with the only exception of a trend towards amelioration in FEV1 at 24 months, which – however – was small and well within the variability of the test. The absence of significant changes in spirometry is not unusual in COPD smokers who stop smoking and particularly in patients with advanced disease and irreversible airway obstruction (44,45), as is the case in our study population. Nonetheless, no worsening in respiratory physiology (including post-bronchodilator FEV1, FVC, and %FEV1/FVC) was reported in COPD patients who were using HTPs.

There are a range of drawbacks to our analysis. First, our results are based on a small cohort of patients with COPD and must be interpreted cautiously. Nonetheless, beneficial results were consistently observed in many COPD health metrics over the entire 3-year period of the study. Second, COPD HTP users in this study could represent a self-selected sample; we cannot rule out that only those who experienced respiratory symptom improvement continued to use HTPs and were ultimately selected for the study. Finally, roughly half of the study participants did not undergo the 6MWD test and this limits the statistical strength of our results.

Risk reduction and harm reversal in the form of cigarette substitution with low-risk products appears to be a promising path. Our study in COPD smokers indicates that long-lasting improvements in respiratory symptoms, exercise tolerance, quality of life, and rate of disease exacerbations are possible in those who have substituted cigarettes for HTPs. In COPD patients who smoke, poor quality of life and unsatisfactory response to medical treatment remain unmet needs. Anything that can improve their symptoms should not be dismissed light-heartedly. Given that many COPD smokers prefer to smoke despite the negative health effects, cigarette substitution should be seen as a valuable solution to the daunting problem of smoking and combustion-free nicotine delivery technologies, including HTPs, should be considered as part of this strategy. Larger prospective studies on the long-term health impacts of heated tobacco products are needed to validate our preliminary findings on the effectiveness, tolerability, and harm reduction potential of these new technologies in COPD and to generate awareness of the respiratory health risks and benefits of smoking substitution with HTP use that are most important to researchers, policy makers and users.

## Data Availability

The data that support the findings of this study are available from the corresponding author, RP, upon reasonable request.

## Authors’ Contributions

RP – study design, data interpretation, manuscript drafting and revision.

JBM – data analyses, data interpretation, manuscript drafting and revision.

UP - review of medical records and data collection.

BB - review of medical records and data collection.

AP - review of medical records and data collection.

GG – data interpretation and manuscript revision.

SR – manuscript revision.

MM - review of medical records and data collection.

PC – study design, data interpretation and manuscript revision.

All authors have read and approved the final manuscript.

